# Sex-specific Metabolic Signatures of Insulin Resistance, Body Mass Index, and Visceral Adiposity in Fasting and Postprandial States

**DOI:** 10.1101/2024.10.03.24314825

**Authors:** Zahra Shojaeifard, Negar Chahibakhsh, David Horner, Ann-Marie M Schoos, Jonathan Thorsen, Jakob Stokholm, Rebecca Vinding, Klaus Bønnelykke, Morten A. Rasmussen, Parvaneh Ebrahimi

## Abstract

The rising prevalence of obesity and insulin resistance, a risk factor for type 2 diabetes (T2D), among adolescents is a growing public health concern. Understanding the metabolic underpinnings of adiposity and insulin resistance in adolescence can provide insights into the development of metabolic dysfunction, and potentially facilitate early intervention strategies to prevent the progression of these conditions into more severe metabolic disorders in adulthood. We explored the metabolic signatures of insulin resistance, Body Mass Index (BMI), and visceral adiposity in adolescents, in both fasting and postprandial states. A meal challenge was undertaken on 18-year-olds (154 females; 144 males), and their blood metabolites were profiled using Nuclear Magnetic Resonance spectroscopy (NMR). Least Absolute Shrinkage and Selection Operator (LASSO) regression was used for modeling and variable selection, in a sex-stratified manner. The results show distinct metabolic patterns between sexes, with males showing more pronounced postprandial responses and stronger associations between blood metabolome and insulin resistance, BMI, and visceral adiposity. Key metabolites such as lipid metabolites, Branched-Chain Amino Acids (BCAAs), glucose, and Glycoprotein Acetylation (GlycA) were selected as important metabolic entities in predicting insulin resistance and adiposity in adolescence. The findings underscore the complex interplay of metabolites with metabolic health and sex, and can pave the way for developing targeted interventions and preventive strategies specifically tailored to adolescents. Such interventions can potentially mitigate the risk of progression to more severe metabolic disorders.

## Introduction

The rising prevalence of insulin resistance and obesity in adolescents is a growing public health concern. Insulin resistance, which is a risk factor for type 2 diabetes and is strongly associated with adiposity, is a critical risk factor for various metabolic and cardiovascular diseases [1]. Adolescence is a crucial developmental period, characterized by significant physiological and hormonal changes. It is therefore particularly vulnerable to the development of insulin resistance and obesity and their associated health risks [2-4]. Adolescents with overweight and obesity during these formative years are at increased risk of continuing to be obese into adulthood and of later developing severe metabolic diseases such as type 2 diabetes (T2D) and cardiovascular diseases at an earlier age. According to the latest World Health Organization (WHO) report, the prevalence of obesity continues to rise globally. In 2022, over 390 million children and adolescents aged 5– 19 years were overweight, with 160 million of these individuals living with obesity. This marks a dramatic increase from 1990 when only 8% of this age group was overweight, and just 2% were obese. This upward trend enhances the concern over obesity among young populations and its associated health risks [5]. Therefore, understanding the metabolic underpinnings of insulin resistance and adiposity during this critical period is imperative.

Insulin resistance and obesity are linked to extensive disruptions in metabolic processes, resulting in significant changes in amino acid and fat metabolism [6]. Insulin resistance, higher body mass index (BMI), and excessive adiposity can alter metabolism by reducing the capacity for glucose uptake in muscle and adipose tissues, leading to altered lipid and amino acid profiles [7]. This metabolic dysfunction can subsequently exacerbate the risk of developing further complications such as cardiovascular diseases and T2D [8]. Therefore, metabolic profiling of biosamples such as plasma, serum, and urine can offer valuable insights into the regulation of physiological processes and the development of diseases. Previous studies have identified various metabolites linked to insulin resistance and BMI in adults. For instance, elevated levels of branched-chain amino acids (BCAAs) and certain lipid metabolites have been consistently associated with insulin resistance and increased adiposity [9]. Furthermore, intervention studies have reported that reducing body weight can result in decreased serum triglycerides and insulin resistance [10]. However, research specifically targeting adolescents is limited. Adolescents have unique metabolic profiles and hormonal milieus compared to adults, necessitating focused studies in this population.

To date, most studies have focused on the effect of anthropometric measurements and insulin resistance in the fasting state. However, fasting and postprandial states represent two distinct metabolic conditions that can influence the levels and activity of various metabolites. The fasting state provides a baseline measure of metabolic function, while the postprandial state reveals metabolic responses to nutrient intake. Postprandial metabolites can serve as independent risk factors for non-communicable diseases and obesity, capturing dynamic metabolic responses that are not evident in the fasting state [11, 12]. Exploring postprandial states can enhance our understanding of the dynamic metabolic processes in adolescents and identify how they relate to insulin resistance and obesity. This will provide insight into the underlying mechanisms of insulin resistance and a more comprehensive view of metabolic health.

This study aimed at identifying metabolic signatures of insulin resistance using homeostatic model assessment for insulin resistance (HOMA-IR), BMI, and waist-to-height ratio (WHtR) at both fasting and postprandial states, from a standardized Oral Glucose Protein Lipid Tolerance Test (OGPLTT) among 298 generally healthy adolescents, in a prospective mother-child cohort [13-15]. WHtR is a more direct measure of central adiposity and visceral fat, which is more closely linked to insulin resistance and T2D than BMI and overall body adiposity [16-18]. The limitation of BMI is that it does not distinguish between muscle and fat or between different types of fat distribution, which can lead to misclassification of individuals’ health risks [16-18]. Based on the sex differences in susceptibility to insulin resistance [19], body composition, and adiposity distribution [20, 21], all our analyses were sex-stratified. We used the least absolute shrinkage and selection operator (LASSO) regression [22] for variable selection and regularization, in order to enhance the interpretability of our statistical models. The LASSO is particularly well-suited for datasets exhibiting high levels of multicollinearity between the variables, and hence is a suitable choice for metabolomic data. Our models identified metabolites that are associated with insulin resistance, BMI, and body adiposity, and pinpoint the differences between fasting and postprandial states, as well as the differences between sexes. Our findings can enhance the understanding of the mechanisms underlying insulin resistance and obesity, paving the way for developing targeted interventions and preventive strategies specifically tailored to adolescents. Ultimately, this could reduce the risk of progression to more severe metabolic disorders.

## Results

An OGPLTT was conducted with 298 18-year-old participants (154 females, 144 males) from the Copenhagen Prospective Studies of Asthma in Children 2000 (COPSAC2000) prospective birth cohort, which originally included 411 infants followed from 1 month of age. Blood samples were collected and analyzed using nuclear magnetic resonance spectroscopy (NMR) to measure blood metabolites at both fasting and postprandial states (0–240 min) in a total of 8 consecutive samples. The experimental design is illustrated in Figure 1, and the characteristics of the participants are detailed in Table 1.

**Fig 1.**
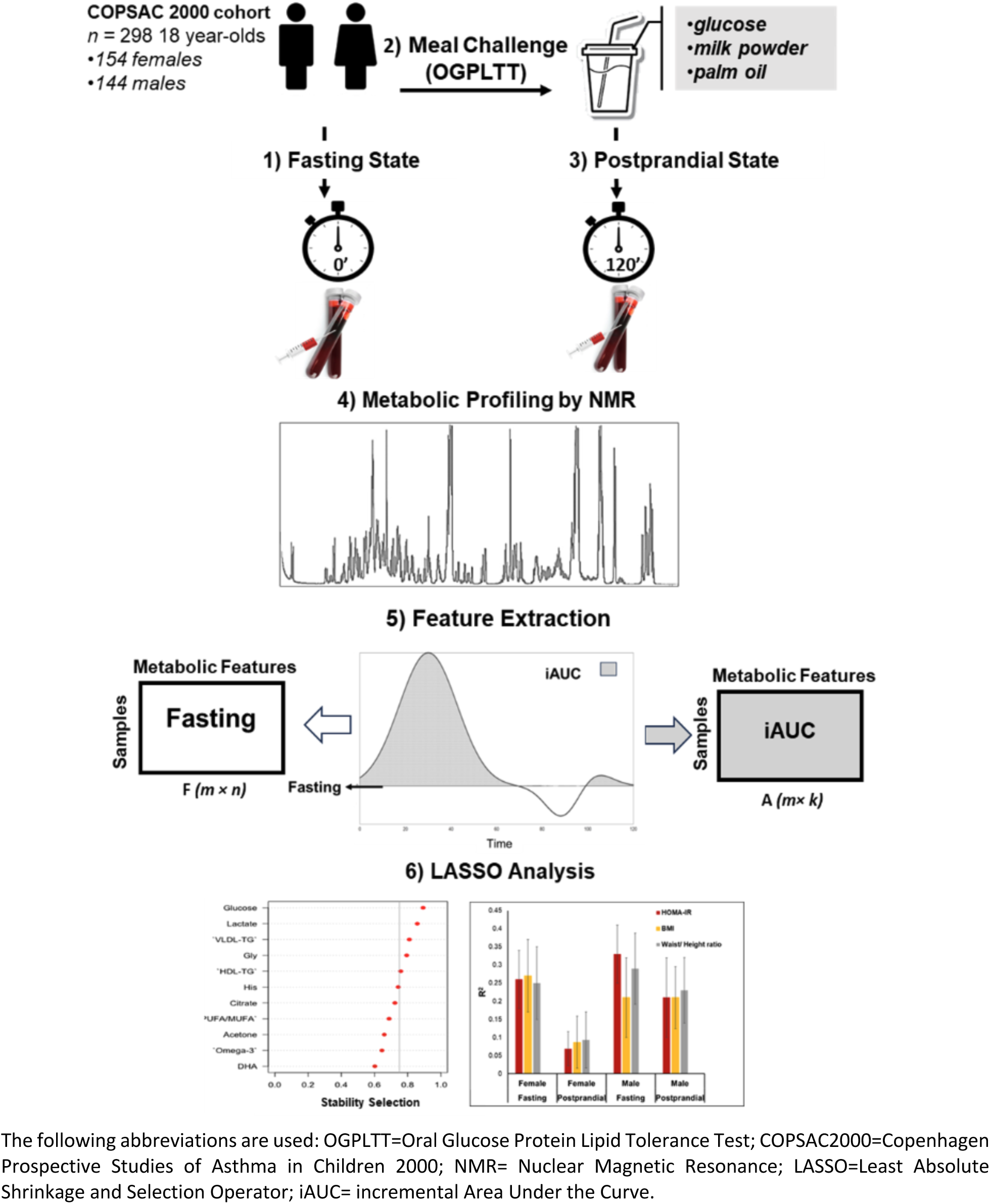
Workflow of the study design. An OGPLTT was conducted, and samples were collected from 298 healthy 18-year-olds (154 females, 144 males) in fasting and postprandial (0–240 min) conditions in the COPSAC2000 study. We analyzed blood metabolites by NMR, and investigated associations of the metabolites using LASSO regression in both sexes, incorporating baseline metabolites from the fasting state and iAUC for the postprandial condition at 120 minutes.

**Table 1.** Descriptive characteristics of the participants in the meal challenge. The first number under each column is the median, and the numbers in the parentheses are the 25th percentile (Q1) and the 75th percentile (Q3), respectively, showing the Inter-Quartile Range (IQR).75th percentile (Q3).

| Characteristic | Overall, N = 298<sup>1</sup> | Female, N = 154<sup>1</sup> | Male, N = 144<sup>1</sup> | p-value<sup>2</sup> |
| --- | --- | --- | --- | --- |
| BMI (kg/m<sup>2</sup>) | 22.2 (19.9, 24.6) | 22.3 (19.8, 24.8) | 22.1 (20.1, 24.2) | 0.8 |
| HOMA-IR | 2.27 (1.54, 3.40) | 2.66 (1.79, 3.69) | 2.02 (1.47, 3.03) | <0.001 |
| Blood glucose (mg/dL) | 97 (92, 103) | 97 (92, 103) | 99 (94, 103) | 0.3 |
| Blood insulin (mU/L) | 9.6 (6.5, 14.0) | 11.2 (7.6, 16.2) | 8.5 (5.9, 12.4) | <0.001 |
| Weight (kg) | 67 (61, 78) | 63 (57, 71) | 73 (66, 81) | <0.001 |
| Height (m) | 1.75 (1.68, 1.81) | 1.68 (1.64, 1.73) | 1.81 (1.77, 1.87) | <0.001 |
| WC (cm) | 78 (73, 84) | 77 (72, 84) | 78 (74, 84) | 0.057 |
| HC (cm) | 87 (82, 95) | 89 (83, 96) | 86 (80, 92) | 0.002 |
| WHR | 0.92 (0.85, 0.96) | 0.88 (0.82, 0.95) | 0.94 (0.91, 0.97) | <0.001 |
| BF (%) | 23 (17, 29) | 28 (25, 32) | 16 (14, 20) | <0.001 |
| MM (kg) | 49 (43, 57) | 43 (40, 46) | 57 (53, 62) | <0.001 |
| MFR | 3.12 (2.31, 4.71) | 2.43 (2.04, 2.86) | 4.83 (3.69, 5.65) | <0.001 |
| WHtR | 44 (41, 48) | 46 (42, 50) | 43 (41, 46) | <0.001 |
| FFM index (kg/m<sup>2</sup>) | 17.1 (15.6, 18.6) | 15.8 (14.9, 16.9) | 18.4 (17.3, 19.4) | <0.001 |
| SBP (mmHg) | 115 (108, 123) | 112 (106, 119) | 120 (114, 126) | <0.001 |
| DBP (mmHg) | 71.0 (67.5, 75.1) | 70.5 (67.5, 74.5) | 71.5 (67.5, 76.0) | 0.3 |
| Social circumstances* | -0.06 (-0.72, 0.67) | 0.08 (-0.61, 0.75) | -0.19 (-0.81, 0.53) | 0.028 |

<sup>1</sup>Median (IQR)
<sup>2</sup>Wilcoxon rank sum test
\* Social circumstances were estimated by PCA using household income, maternal education, and mother's age when the child was 2 years old.
The following abbreviations are used: BMI = Body Mass Index; HOMA-IR = Homeostasis Model Assessment Insulin Resistance; WC= Waist Circumference; HC= Hip Circumference; WHR= Waist-to-Hip Ratio; %BF= Body Fat Percentage; MM= Muscle Mass; MFR= Muscle-to-Fat Ratio; WHtR= Waits-to-Height Ratio; FFM= Fat-free-Mass index; SBP=Systolic Blood Pressure; DBP= Diastolic Blood Pressure.

HOMA-IR and WHtR were higher in females than in males (*P* < 10^-3^), while BMI was not significantly different (Table 1). Linear models were fitted between HOMA-IR and BMI (Supplementary Fig. 1). As expected, there was a significant positive association between BMI and HOMA-IR in both sexes, where almost one-third of the variance in HOMA-IR was explained by BMI (*R*^2^ ≈ 0.3; *P* < 10^-3^), indicating that other factors also contribute to HOMA-IR variability.

We investigated the associations between fasting and postprandial blood metabolites with HOMA-IR, BMI, and WHtR, using LASSO regression in sex-stratified analyses. For the fasting state, the baseline metabolite concentrations were used, while for the postprandial state, the incremental area under the curve (iAUC) of the metabolic profiles at 120 min was calculated. Fig. 2 and Supplementary Table 1 show the statistics of the LASSO models. The models between HOMA-IR and fasting metabolites revealed differences in predictive performance between males and females. The model for males was stronger, explaining approximately 32.6% of the variance in HOMA-IR, compared to 25.8% in females. In the postprandial stage, the difference between the sexes became more pronounced; the model for males, using the iAUC of metabolites at 120 min, explained 21.0% of the variance in HOMA-IR, while the model for females accounted for only 7.0%.

**Fig 2.**
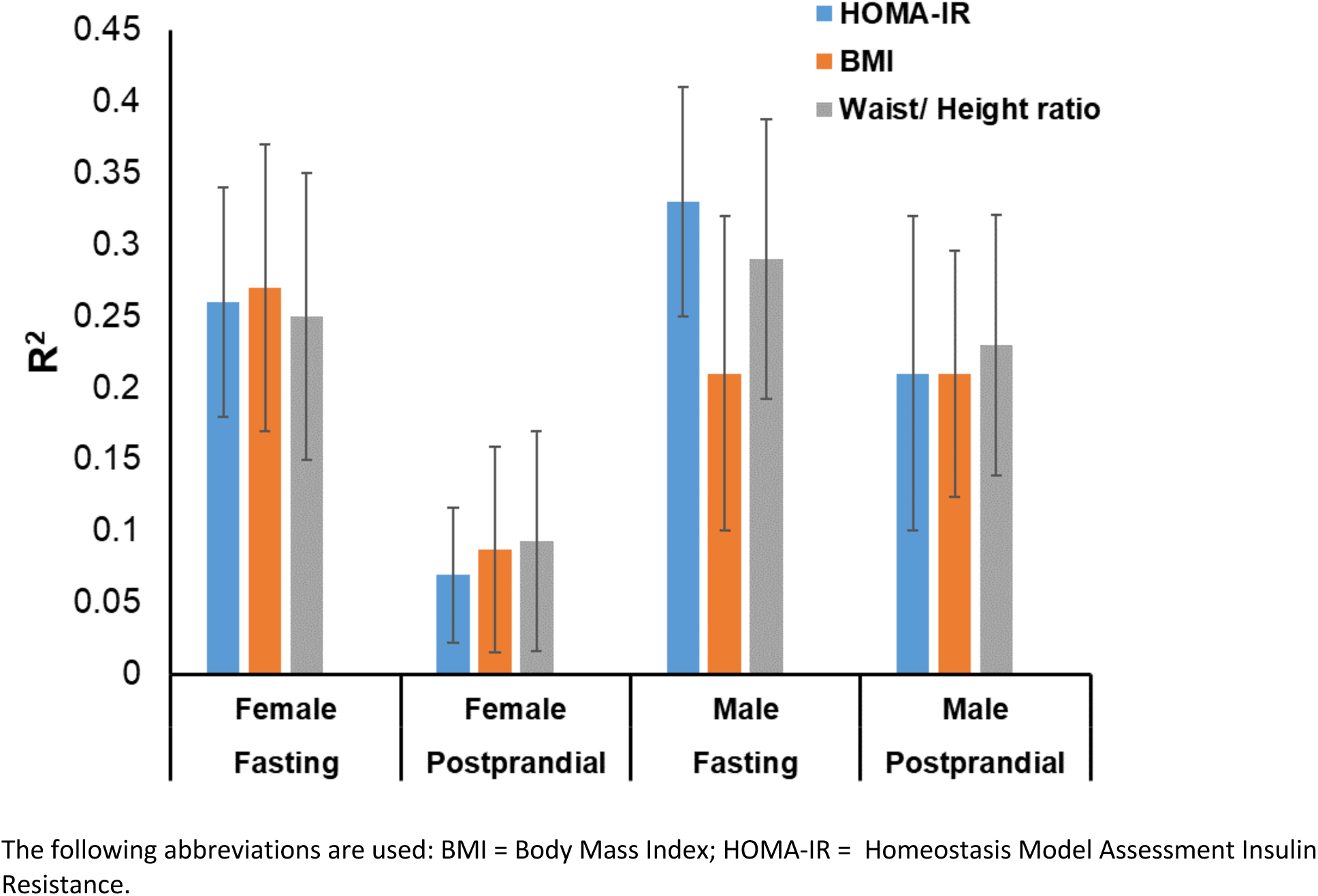
Summary of the LASSO models. The bars show the standard deviation of the coefficient of determination (R²).

The models assessing the relationship between BMI and fasting metabolites demonstrated some variation in predictive performance between males and females. The model for females explained 26.8% of the variance in BMI, while the model for males explained slightly less, at 21.0%. In the postprandial stage, the variation between males and females was more pronounced. Again, the model for males exhibited a stronger explanatory power, capturing 20.8% of the variance in BMI, compared to just 8.7% in females.

The models between WHtR and fasting metabolites showed slightly better fit and predictive accuracy in males than in females. The models explained around 24% and 29% of the variation in WHtR in females and males, respectively. A similar pattern was observed in the postprandial state, with male models explaining 23% of the variance and female models explaining only 9% (See Fig. 2 and Supplementary Table 1).

To identify the most stable LASSO features and predictors, we employed stability selection, iterating the model training process to reliably pinpoint the most robust and consistent features. Fig. 3 and 4 show the LASSO coefficient of stable features with a selection probability of ≥ 0.75 over 500 iterations, and Supplementary Fig. 2A and 2B show the stability selection plots, separately for fasting and postprandial states.

**Fig 3.**
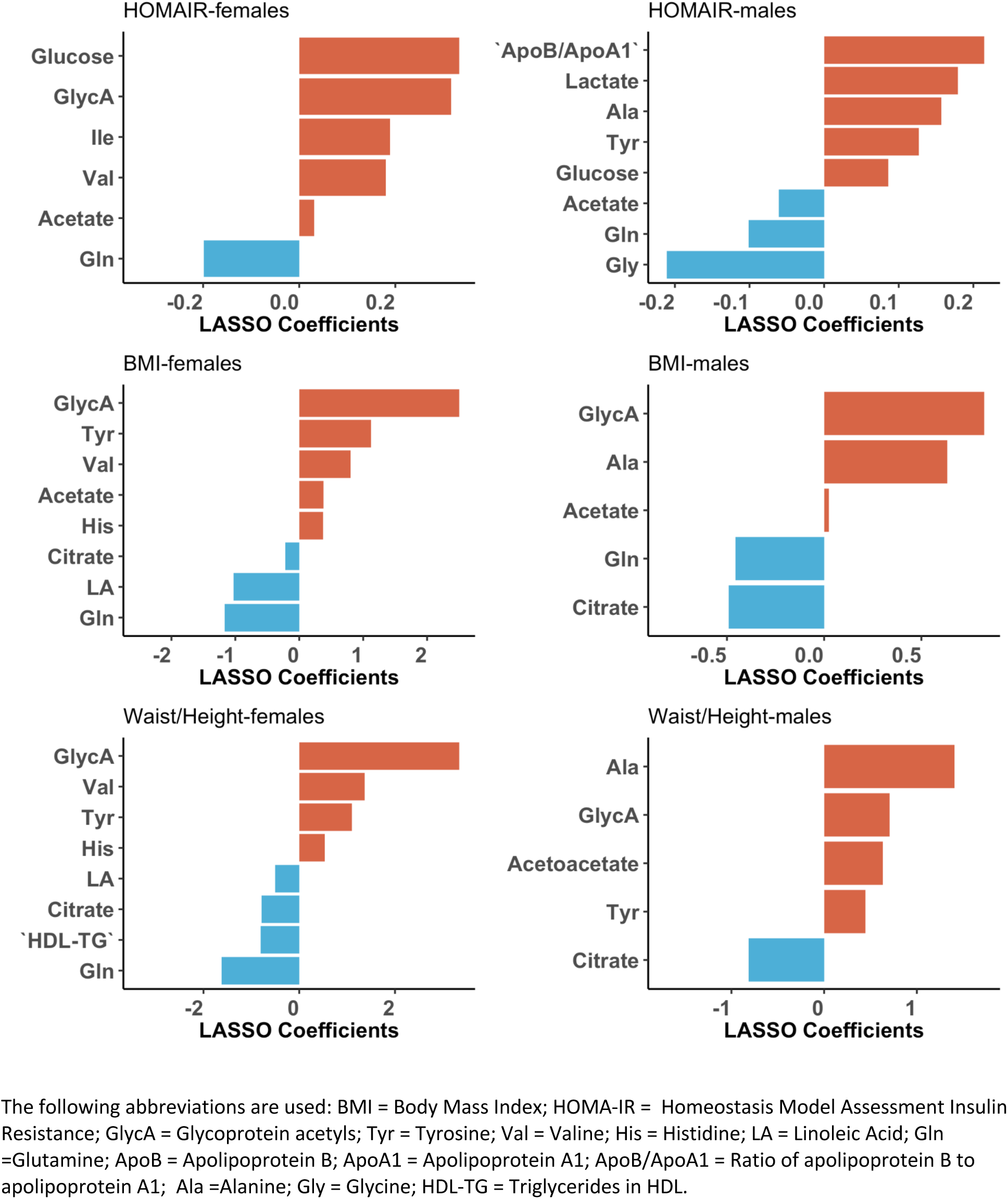
LASSO coefficient of stable features with selection probability of ≥0.75 during 500 subsampling replicates, at fasting state.

**Fig 4.**
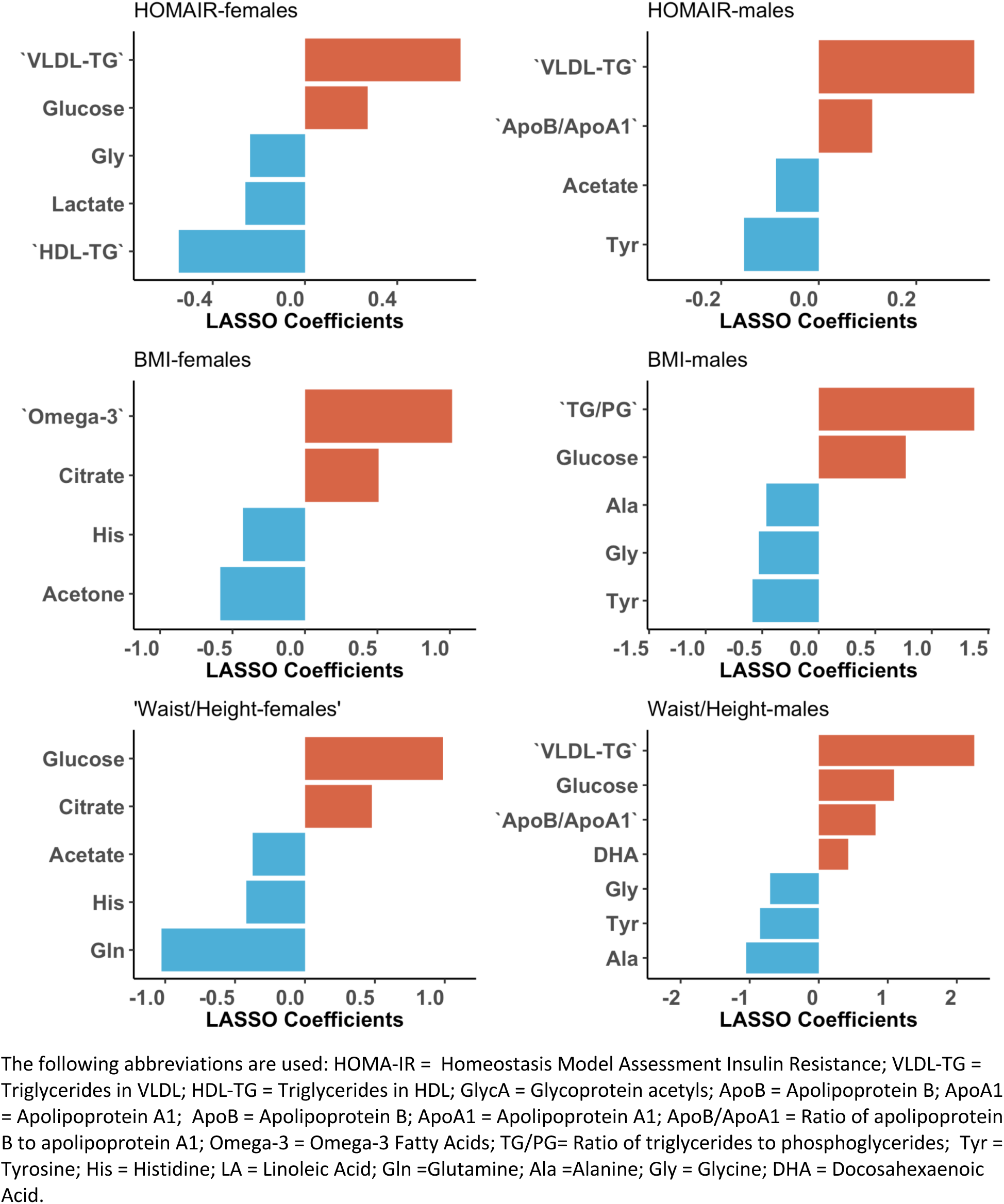
LASSO coefficient of stable features with selection probability of ≥0.75 during 500 subsampling replicates, at postprandial state.

*Fasting state:* For HOMA-IR in the fasting state, positive associations in females were observed with glucose, glycoprotein acetyls (GlycA), isoleucine (Ile), valine (Val), and acetate, while glutamine (Gln) showed a negative association. In males, HOMA-IR was positively associated with the ratio of Apolipoprotein B to Apolipoprotein A1 (ApoB/ApoA1), lactate, alanine (Ala), tyrosine (Tyr), and glucose, whereas acetate, Gln, and glycine (Gly) were negatively associated. Regarding BMI in the fasting state, positive associations in females included GlycA, Tyr, Val, acetate, and histidine (His), while negative associations were observed for citrate, linoleic acid (LA), and Gln. In males, BMI was positively associated with GlycA, Ala, and acetate, and negatively associated with Gln and citrate. For WHtR in the fasting state, GlycA, Val, Tyr, and His showed a positive association in females, whereas LA, citrate, high-density lipoprotein triglycerides (HDL-TG), and Gln showed a negative association. In males, positive coefficients included Ala, GlycA, acetoacetate, and Tyr, whereas citrate was the only stable feature with a negative coefficient (See Fig. 3).

*Postprandial state:* For HOMA-IR in the postprandial state, positive associations in females were observed with very-low-density lipoprotein triglycerides (VLDL-TG) and glucose, while negative associations were noted with Gly, lactate, and HDL-TG. In males, HOMA-IR was positively associated with VLDL-TG and ApoB/ApoA1, and negatively associated with acetate and Tyr. Regarding BMI in the postprandial state, positive associations in females included Omega-3 and citrate, while negative associations were found with His, and acetone. In males, BMI was positively associated with triglycerides to phosphoglycerides ratio (TG/PG) and glucose, and negatively associated with Ala, Gly, and Tyr. For WHtR in the postprandial state, glucose and citrate showed positive associations in females, whereas acetate, His, and Gln were negatively associated. In males, VLDL-TG, glucose, ApoB/ApoA1, and docosahexaenoic acid (DHA) showed positive associations, whereas Gly, Tyr, and Ala had negative coefficients (See Fig. 4).

Principal Component Analysis (PCA) was performed on both fasting and postprandial datasets, following subsetting based on the stable LASSO-selected features for HOMA-IR, BMI, and WHtR (as shown in Fig. 3 and Fig. 4). PCA biplots are presented in Supplementary Fig. 3, where the PCA scores are colored according to HOMA-IR, BMI, and WHtR categories. HOMA-IR and BMI are dichotomized into high and low groups using the thresholds of 24.9 for BMI and 2.5 for HOMA-IR [23, 24]. For WHtR, the median in females and males was used as the cutoff value for high and low. In models with comparatively better fit (See Fig. 2 and Supplementary Table 1), such as HOMA-IR versus fasting metabolome in both males and females, a moderate clustering of the samples is evident within the first two principal components. Conversely, in models with weaker fit, such as BMI versus postprandial metabolome in females, no distinct clustering patterns are observed.

## Discussion

Our findings revealed distinct metabolic signatures associated with insulin resistance, BMI, and WHtR in adolescents during both fasting and at the 120-min postprandial time point. The rationale for selecting 120 minutes as the postprandial time point of interest is that glucose and insulin levels typically peak around 2 hours after eating. Therefore, focusing on this time point captures the most significant changes in metabolite concentrations and reflects the body’s immediate metabolic response [25]. Consistent with our hypothesis, the necessity of a sex-stratified analysis was evident, as the observed differences between the female and male models clearly reflected known biological variations in metabolic responses and body composition among the sexes. This once again underscores the importance of considering sex-specific factors in metabolic research.

In general, the models showed better fit in males, both at fasting and postprandial states. This was more pronounced at the postprandial state, where HOMA-IR, BMI, and WHtR were associated with much stronger postprandial responses in males than in females. In females, insulin resistance and anthropometric measures reflected mainly at the fasting state, and the models had poor fit at the postprandial state. However, in males, the models at the postprandial state were comparable to the fasting state. These indicate that males experience more pronounced metabolic shifts in response to food intake, where insulin resistance, BMI, and WHtR were more closely linked to the postprandial metabolic changes in males. This is despite the fact that insulin resistance and WHtR were significantly higher in females, which may indicate that males may be more prone to deterioration in metabolic health due to insulin resistance and higher body adiposity.

In females, the BMI and WHtR models had similar fit at the fasting state, but in males, WHtR was reflected better than BMI. Considering that BMI is a measure of overall body composition and WHtR is a measure of metabolically active visceral fat, these observations indicate that in males, abdominal fat plays a more significant role in influencing the metabolome than total body fat. Previous studies have shown that visceral fat in male adolescents is associated more strongly with adiposity-related conditions (such as hypertension) than BMI is, which was not the case in females [26].

The features selected by LASSO across different models provide valuable insights into the key metabolic patterns and alterations associated with insulin resistance, BMI, and visceral fat. Considering the poor fit of the postprandial models in females, selected metabolites from these models are not discussed. The positive coefficients observed for glucose and GlycA in several of the models align with previous findings suggesting their role in insulin resistance and inflammation [27, 28].

Lipid metabolism markers were represented more in males. VLDL-TG showed positive associations with HOMA-IR and WHtR at the postprandial state in males, consistent with studies associating postprandial hyperlipidemia with insulin resistance and metabolic syndrome [29, 30]. In males, ApoB/ApoA1 also showed a positive association with HOMA-IR at both fasting and postprandial states, and with WHtR postprandially. ApoA1 is the principal apolipoprotein associated with high-density lipoprotein (HDL), playing a crucial role in mediating the antioxidant and anti-inflammatory properties of HDL. In contrast, ApoB is a key structural component of very low-density lipoproteins (VLDLs) and their metabolites. The ApoB/ApoA1 ratio, therefore, serves as an important indicator of the balance between potentially atherogenic and anti-atherogenic lipoprotein particles. A higher ratio is positively correlated with an increased risk of atherogenesis and cardiovascular disease, reflecting a shift towards a more atherogenic lipid profile [31]. Based on these, the positive association that we observed in our cohort between the ApoB/ApoA1 ratio and HOMA-IR and WHtR in males suggests that even at 18 years of age, insulin resistance and visceral fat are beginning to exert detrimental effects on metabolic health, potentially increasing the risk of severe metabolic and cardiovascular issues later in adulthood. The TG/PG ratio was one of the other lipid metabolism-related features selected that was positively associated with BMI in males at the postprandial state. The TG/PG ratio has been previously associated with metabolic syndrome and metabolically unhealthy overweight [32].

Ile and Val appear in several of the models, showing a positive association with insulin resistance and adiposity. There is substantial evidence linking elevated levels of BCAA with obesity, insulin resistance, and type 2 diabetes [33]. The relationship between BCAAs and metabolic health is complex, with some studies suggesting that increased BCAA levels may be both a cause and consequence of metabolic disturbances [34]. From the amino acids group, Tyr and Ala also had positive coefficients in the fasting state in several of the models in both sexes, which aligns with the literature where elevated levels of these amino acids in the blood have been associated with insulin resistance and higher body adiposity [35, 36]. On the other hand, Ala and Tyr showed negative associations in the male models at the postprandial state, meaning a lower increase of these metabolites after the meal-intake relative to the baseline. Such divergences in associations between the fasting and postprandial states for some of the metabolites highlight the complex and dynamic nature of metabolic regulations. Two other amino acids showing negative associations with insulin resistance and adiposity were Gln and Gly, where Gly was only selected in the male models. These amino acids have previously been negatively associated with obesity [37, 38], and glutamine supplementation has been shown to reduce BMI and improve insulin resistance [39].

Citrate consistently appeared in the BMI and WHtR models at the fasting state, with negative coefficients for these measures in both sexes. This is in line with studies that have reported an inverse relation between plasma citrate levels and BMI [40, 41]. Lactate was selected only in the insulin resistance model for males at the fasting state, with positive association. However, considering that it was one of the most robust features with a high coefficient, it is being briefly discussed. Plasma lactate has been previously reported as a useful marker of metabolic health and insulin resistance, and elevated lactate levels can indicate increased reliance on glycolysis and reduced insulin sensitivity [42]. Another interesting feature was LA which showed a negative association with the adiposity measures in females at the fasting state. Previous studies have reported that supplementation with LA can slightly reduce BMI [43], and it has also been reported to have anti-diabetic effects [44].

Our study identified significant associations between blood metabolites and HOMA-IR, BMI, and WHtR in both fasting and postprandial states, with distinct differences between sexes, highlighting metabolites underlying insulin resistance and obesity among adolescents. The findings underscore the complex interplay of metabolites with metabolic health, fasting and postprandial metabolic states, and sex, and can pave the way for developing targeted interventions and preventive strategies, specifically tailored to adolescents. Ultimately, this could reduce the risk of progression to more severe metabolic disorders.

Most studies on metabolic health have been conducted on adult cohorts, whereas investigating the effects of anthropometric measurements and insulin resistance in adolescents provides an opportunity to study these factors in individuals who are still relatively healthy and free from concurrent morbidities. Adolescents represent a unique demographic where early intervention could prevent the progression of insulin resistance and adiposity into more severe health conditions later in life. Moreover, we had a sex-balanced design, and by performing a sex-stratified analysis, we highlighted potential sex-specific signatures of insulin resistance and BMI. The other advantage of our study was investigating not only the fasting state, but also carrying out a standardized diet challenge and investigating the postprandial response to understand how it is regulated by insulin sensitivity and body adiposity.

Future research could build on our findings to explore the longitudinal impact of these metabolic signatures and their potential as biomarkers for early detection and intervention. In addition, further research could explore the mechanisms underlying the identified associations and their implications for metabolic disease prevention and treatment.

## Methods

### Study population

The participants were part of the COPSAC2000 cohort, which is a prospective clinical mother-child cohort study of 411 children of asthmatic mothers [13, 14]. The participants were deeply phenotyped with 17 scheduled visits to the COPSAC research clinic from birth till age 18. At the 18-year visit, 298 of the participants underwent an Oral Glucose Protein Lipid Tolerance Test (OGPLTT).

### The meal challenge and blood sample collection

298 (73%) out of the 411 participants in the COPSAC2000 cohort underwent and completed a meal challenge at the 18-year visit [15]. There were various reasons for the dropouts, including vomiting, fear of needles, failing to insert intravenous line, lactose intolerance, vegan diet, diabetes or other diseases, or plain refusal to participate in this part of the 18-year visit. The participants had fasted for at least 8 hours before the challenge. After the insertion of an intravenous catheter and collecting baseline (fasting) blood samples, they received an OGPLTT. The meal was a hot beverage, consisting of 75 g glucose, 60 g palm oil, and 20 g skimmed milk powder, adding up to a total caloric content of ∼947 kcal. This corresponded to ∼33% of the daily caloric expenditure of the participants (∼ 3000 kcal), adjusted for gender and height. Pure vanilla powder was added to improve the taste, and 1.5 g of paracetamol was added to assess ventricular passage time. The meal was consumed within a maximum of 30 min, and preferably 15 min. Once half of the meal was consumed, the timer was set for blood draws at 15, 30, 60, 90, 120, 150, and 240 min postprandially. The test meal contained fat (∼35%), glucose (∼44%), and protein (∼20%).

### Metabolic profiling and extracting metabolic features

Circulating levels of serum metabolites were measured using a targeted high-throughput nuclear magnetic resonance (NMR) metabolomics platform (Nightingale Health Ltd., Helsinki, Finland) for the blood samples from the fasting state and 2h postprandially. From the quantified and calculated metabolic measures from Nightingale, the following categories were excluded before further analysis: lipoprotein subclasses, relative lipoprotein lipid concentrations, fluid balance, and lipoprotein particle sizes.

To extract postprandial features for the remaining metabolites, incremental areas under the curve (iAUC) of the metabolic profiles were calculated [45], using mainly basic R commands and the ‘ggplot2’ package. First, smoothed curves were fitted to individual profiles using all postprandial timepoints. Next, fasting (baseline) levels of the metabolites were subtracted from the fitted curves to get the response specifically during the postprandial period. The iAUC was then calculated by summing the positive increments. For the fasting state, metabolite concentrations in blood samples collected at baseline and before the meal challenge were used.

### BMI, waist-to-height ratio, and insulin resistance

Anthropometrics were assessed during the 18-years visit. Waist circumference was measured using a tape, placing it around the navel as a reference point and averaging two measurements taken during inspiration and expiration. Height was gauged using a stadiometer, calibrated annually, and weight was recorded without clothes and barefoot on calibrated digital scales. Body mass index (BMI) was calculated as body weight divided by body height squared (kg/m2), and waist-to-height ratio (WHtR) was calculated by dividing the waist circumference (cm) by the height (cm). Hip circumference was determined by measuring the widest girth around the hip or buttock area of each participant (cm).

To estimate insulin resistance, homeostatic model assessment for insulin resistance (HOMA-IR) index was used [46], based on the fasting glucose and insulin levels. Fasting glucose and insulin levels in the blood were measured using conventional clinical biochemistry tests. The glucose analysis was conducted using enzymatic determination and absorption photometry, by Siemens Atellica CH 930 equipment. For insulin, sandwich immunoassays using chemiluminescence technology on the Siemens Atellica IM Analyzer were used.

### Data analysis

All the statistical analyses were performed in the R environment, version 3.5.1. Least Absolute Shrinkage and Selection Operator (LASSO) models were fitted between the extracted metabolic features (the fasting levels and iAUC_2h_ of the metabolites) and BMI, WHtR, and HOMA-IR, using the ‘Caret’ package in R. The metabolic features were preprocessed by centering and scaling before the analysis. To identify stable and influential metabolites stability selection was performed using the ‘stabs’ package, with 500 subsampling replicates.

## Data Availability

All data produced in the present study are available upon reasonable request to the authors.

## Ethical approval

The study was approved by the Region Hovedstadens in Denmark (H-16040846) and the Danish Data Protection Agency (2015-41-3696).

## Funding statement

COPSAC is supported by both private and public research funds, all of which are detailed on our website, www.copsac.com. Core support has been provided by the Lundbeck Foundation, the Danish Ministry of Health, the Danish Council for Strategic Research, and the Capital Region Research Foundation. No pharmaceutical companies were involved in this study. The funding agencies had no role in the design or conduct of the study, nor in the collection, management, interpretation of the data, or in the preparation, review, or approval of the manuscript. Additionally, MAR received funding from the Novo Nordisk Foundation (NNF21OC0068517).

## Disclosure of conflict of interest

The authors have completed the ICMJE uniform disclosure form at www.icmje.org/coi_disclosure.pdf and declare: no support from any organization for the submitted work; no financial relationships with any organizations that might have an interest in the submitted work in the previous three years; and no other relationships or activities that could appear to have influenced the submitted work. The data and samples used in this study were collected by the Copenhagen Prospective Studies on Asthma in Childhood (COPSAC), with full acknowledgment of all contributors involved.

## Supplementary material

**Fig. S1.**
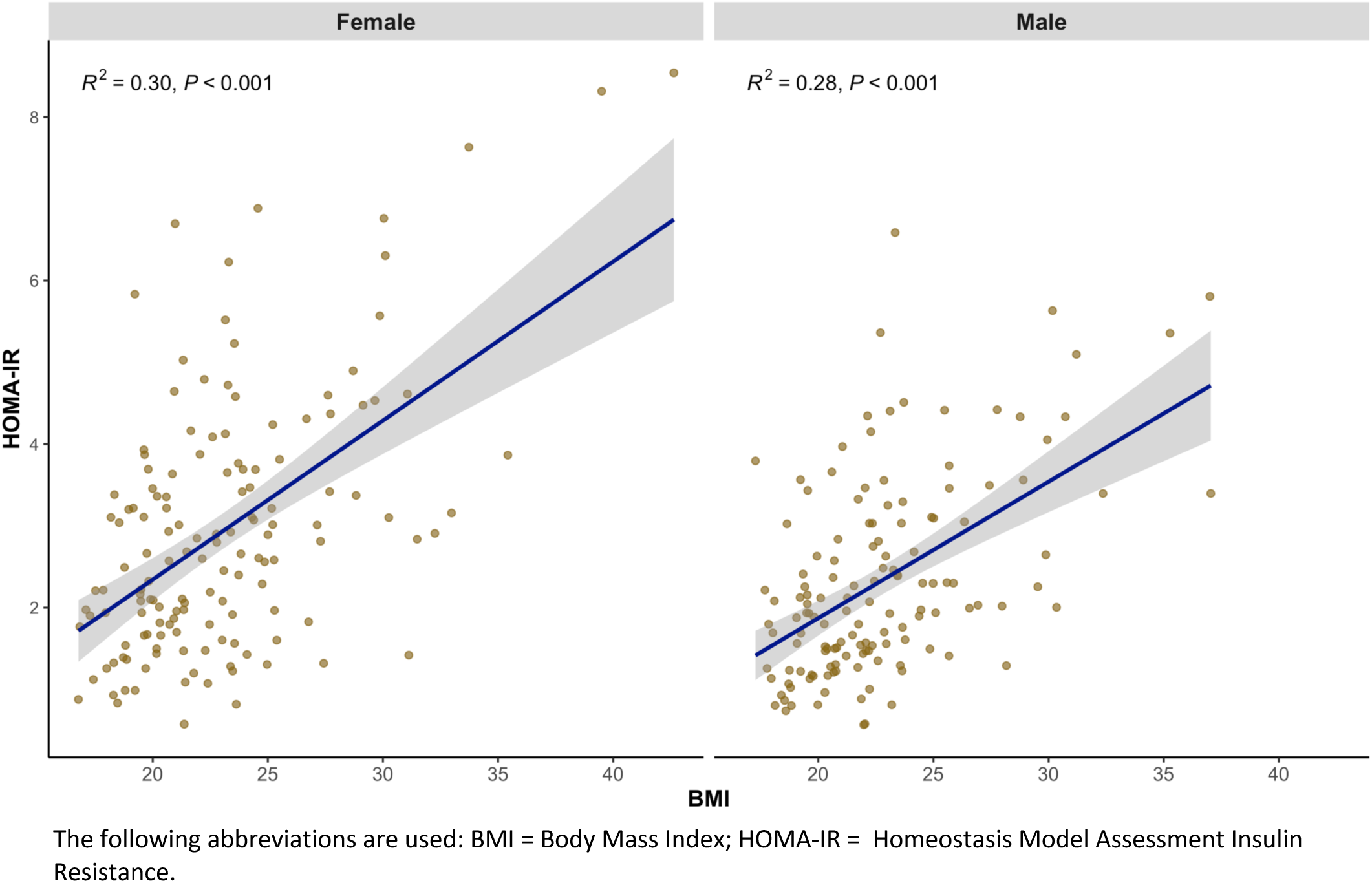
Linear regression analysis of the association between HOMA-IR and BMI.

**Fig. S2.**
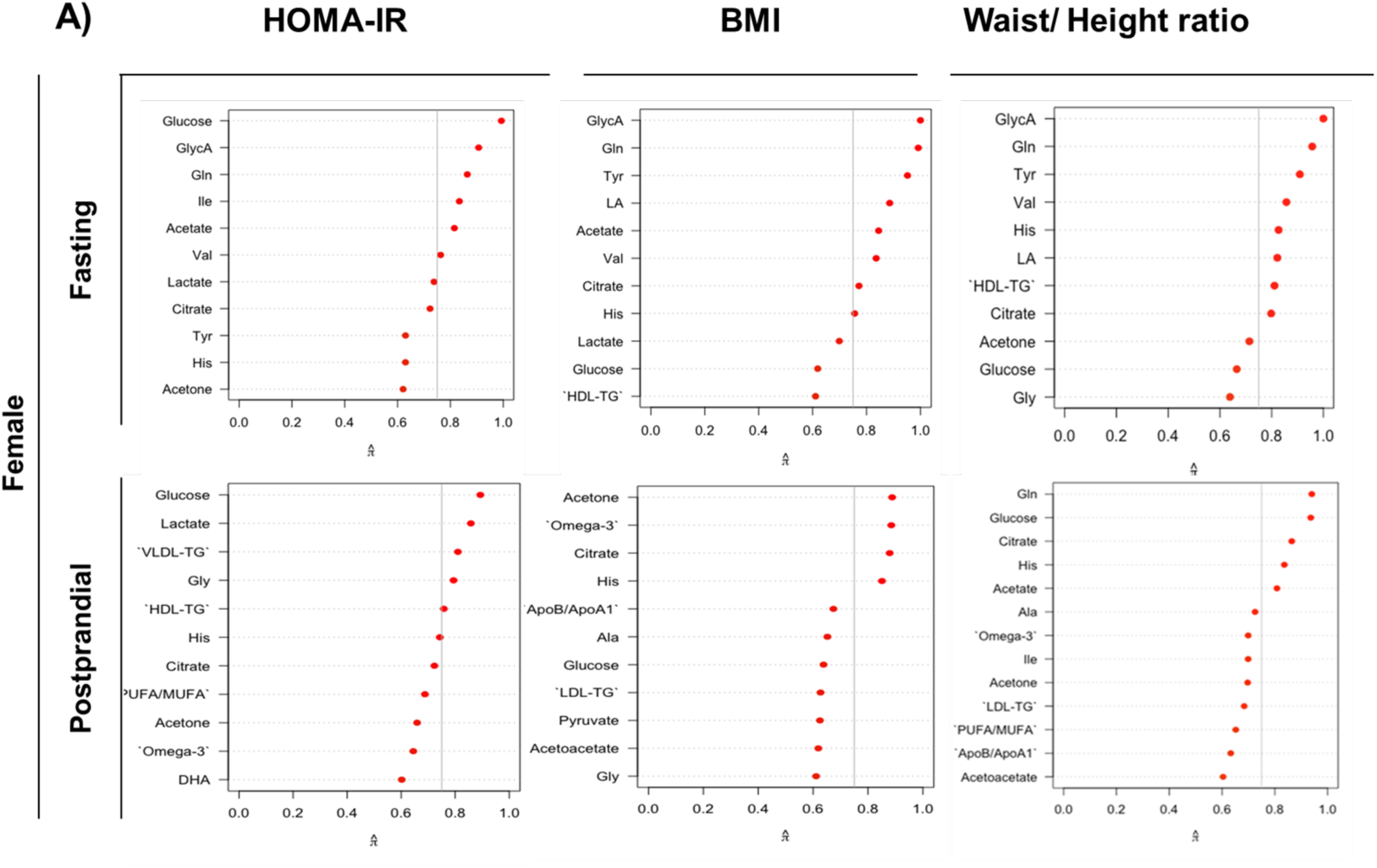
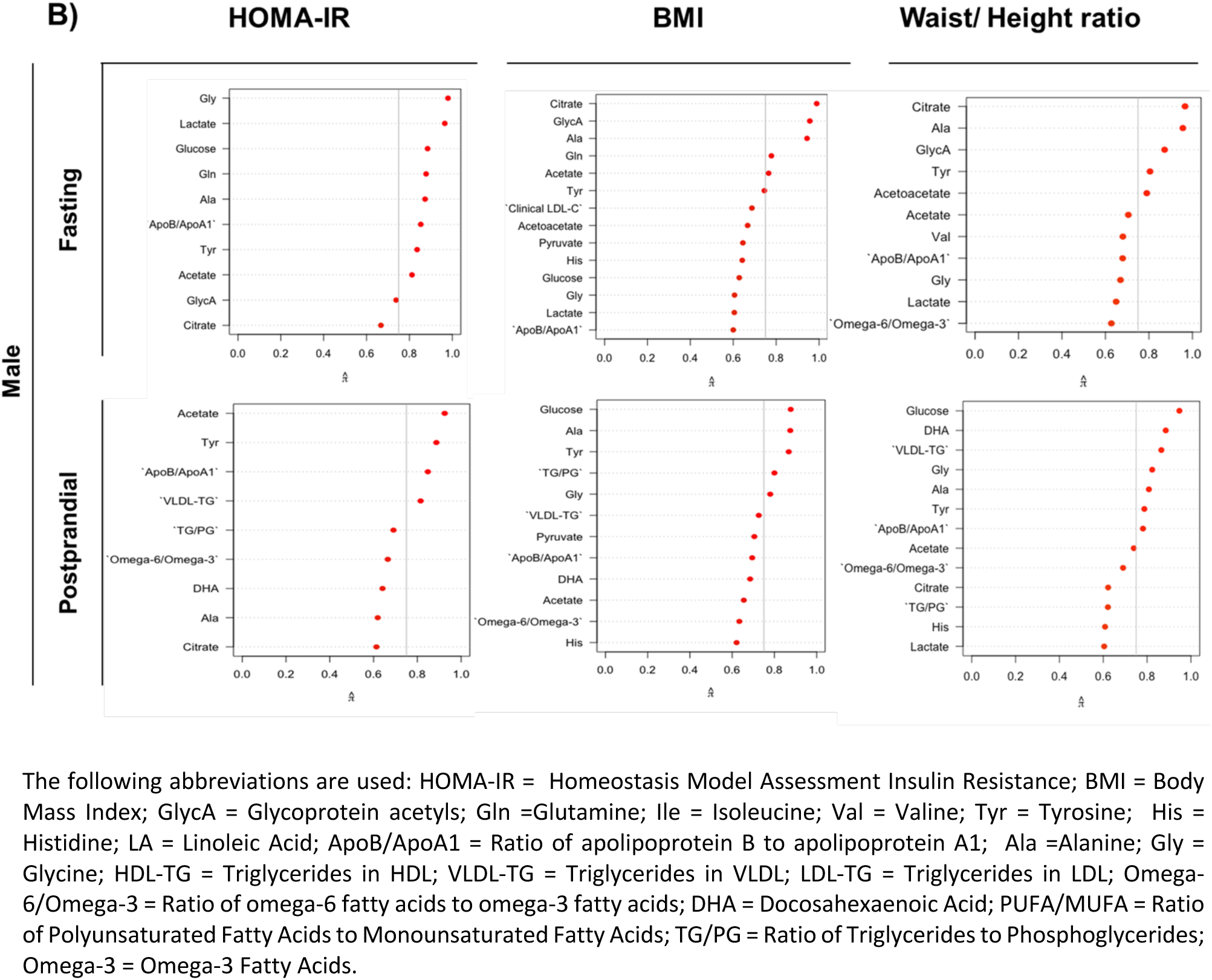
Stability selection plots for the LASSO models, across 500 iterations. 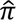 refers to the selection probability or proportion of times a particular feature was selected by the LASSO model across the resampling iterations.

**Table S1.**
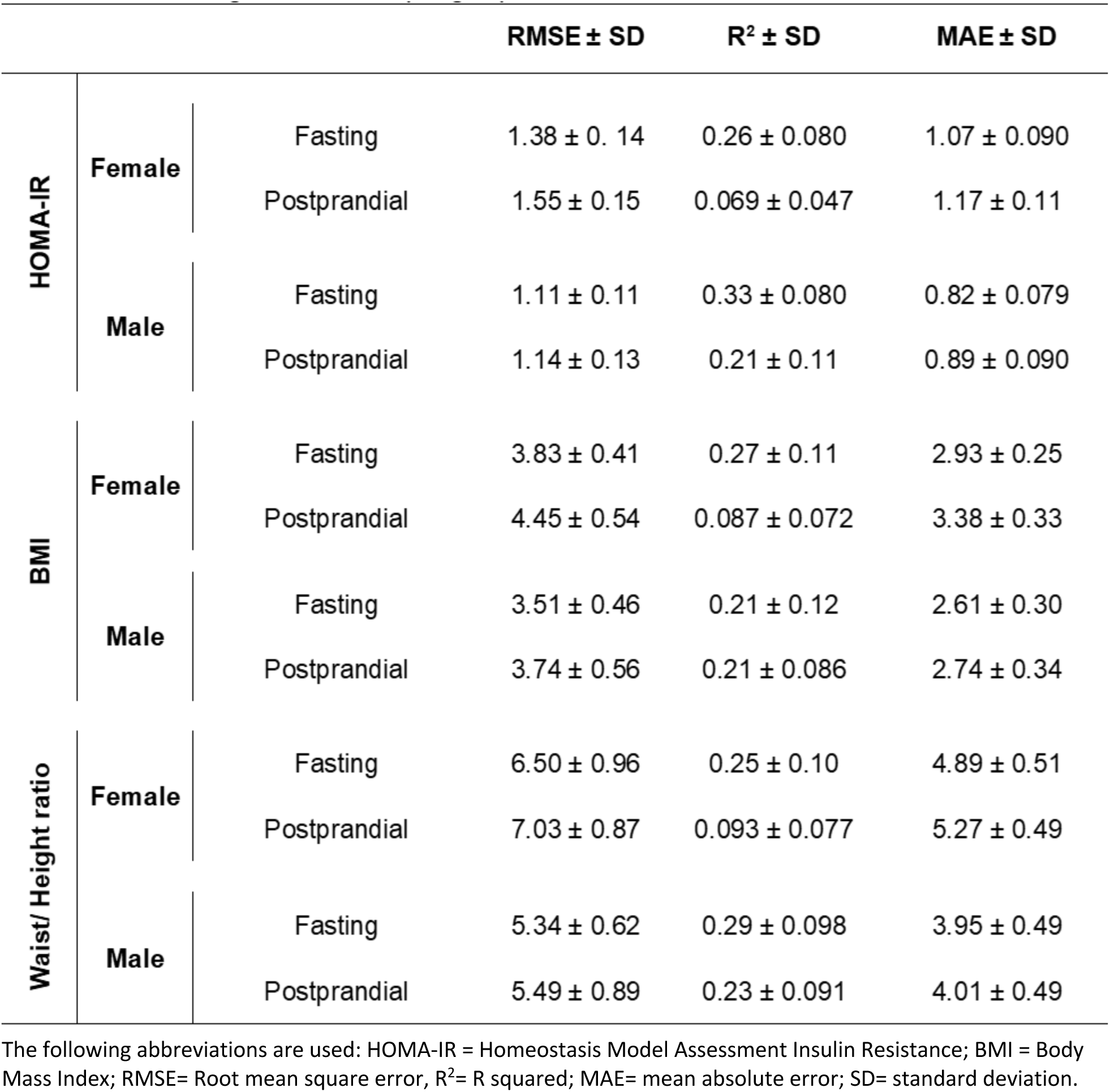
LASSO statistics parameters of HOMA-IR, BMI and Waist/Height ratio metabolites, using 500 subsampling replicates.

**Fig. S3.**
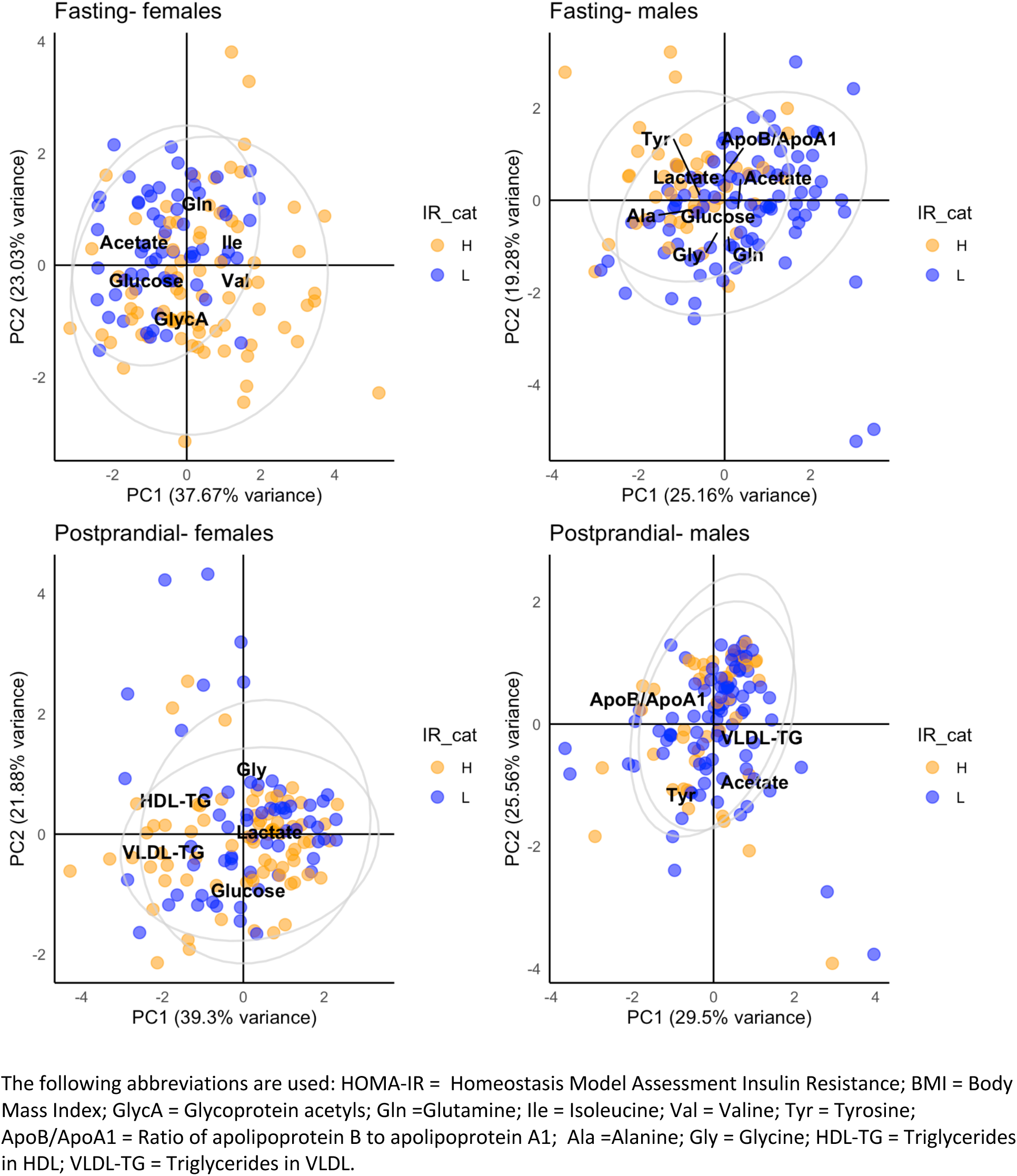
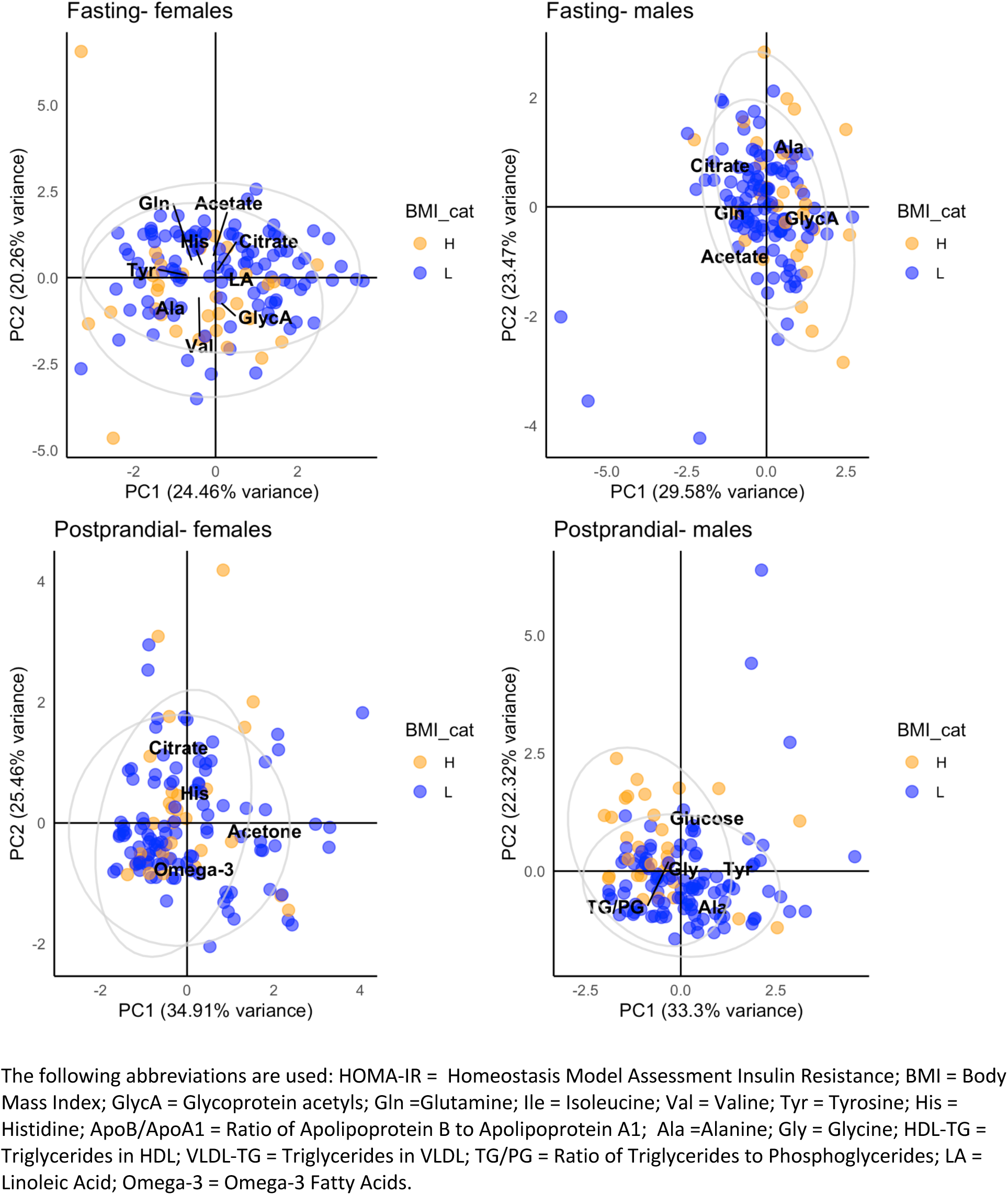
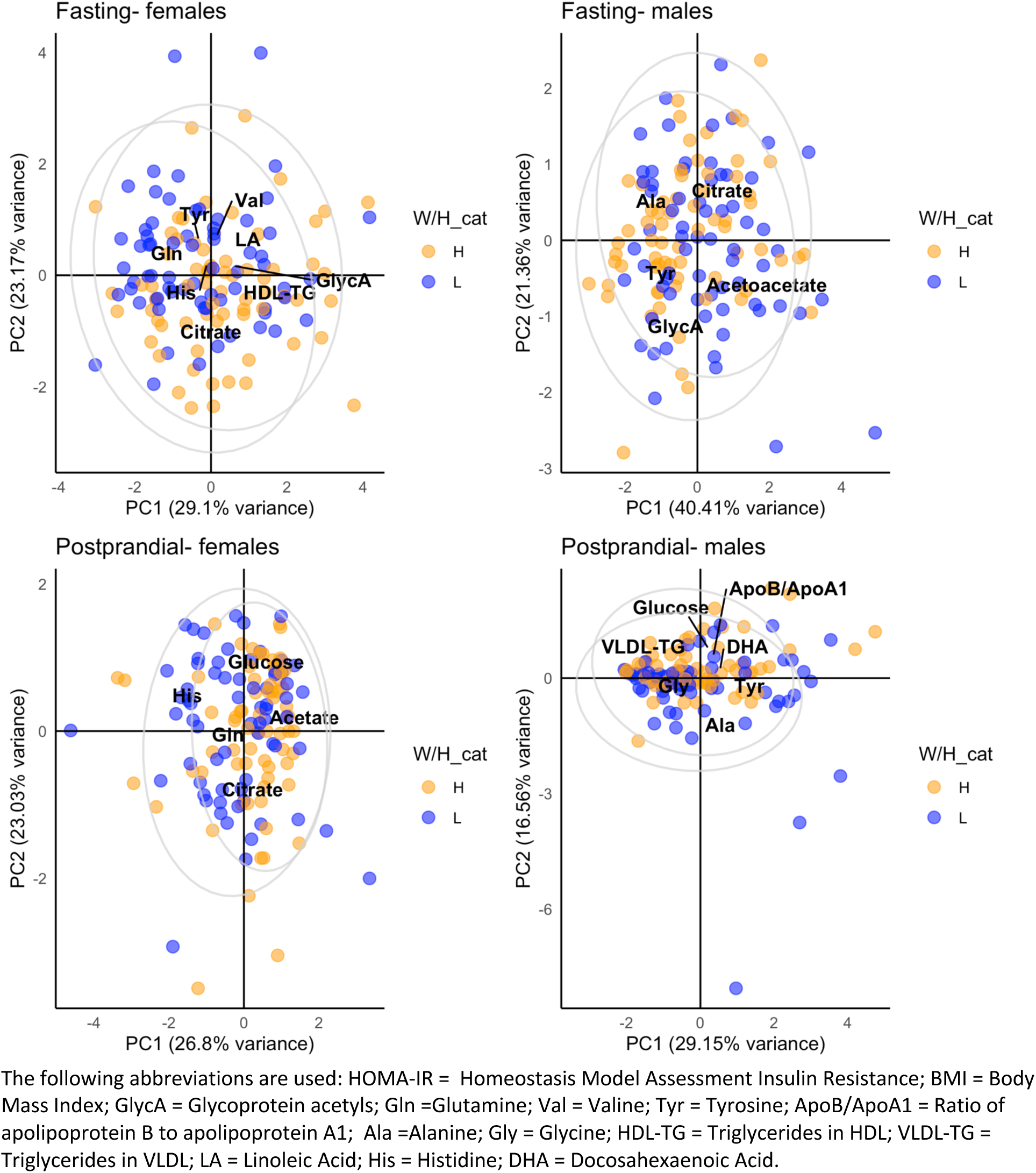
PCA biplots based on the stable LASSO features for BMI and HOMA-IR (listed in Fig. 3 and Fig. 4). The scores are colored by HOMA-IR, BMI, and WHtR categories, dichotomized by the following thresholds as high and low: 24.9 for BMI and 2.5 for HOMA-IR; For WHtR, the median in females and males was used as the cutoff value.

## Notes

### Competing Interest Statement

The authors have declared no competing interest.

### Summary of Updates

Compared with the first revision, two changes have been made: 1) the order of the authors has been corrected 2) the margin of the pages with illustrations has been increased, to prevent overlap with the headings.

